# Hemodynamics After Fontan Procedure are Determined by Patient Characteristics and Anastomosis Placement Not Graft Selection: a Patient-Specific Multiscale Computational Study

**DOI:** 10.1101/2021.10.03.21264033

**Authors:** Ethan Kung, Catriona Baker, Chiara Corsini, Alessia Baretta, Giovanni Biglino, Gregory Arbia, Sanjay Pant, Alison Marsden, Andrew Taylor, Michael Quail, Irene Vignon-Clementel, Giancarlo Pennati, Francesco Migliavacca, Silvia Schievano, Anthony Hlavacek, Adam Dorfman, Tain-Yen Hsia, Richard Figliola, Modeling of Congenital Hearts Alliance (MOCHA)^+^ Investigators

**Author notes:** **Correspondence** Ethan Kung, PhD.

## Abstract

**Objectives:** Patient-specific multiscale modeling simulates virtual surgeries of the Fontan procedure using three different graft options. Predictive modeling details post-operative outcomes that can help inform clinical decision support.

**Methods:** Six patients underwent preoperative cardiac magnetic resonance imaging and catheterization. Virtual surgery is carried out for each patient to test the resulting hemodynamics of three Fontan graft options: ECC, 9mm Y-graft, and 12mm Y-graft.

**Results:** 1) one-way ANOVA p>0.998 in all systemic pressures and flows between graft options, 2) p=0.706 for hepatic flow distribution between graft options, 3) local power loss differences do not affect the systemic circulation, 4) anastomosis positioning modification of the same Y-graft in the same patient changed left PA hepatic distribution from 0.66 to 0.49

**Conclusions:** Systemic pressures and blood flow after the Fontan procedure are not affected by graft selection but are well influenced by patient pulmonary vascular impedance. The hepatic distribution can be affected by anastomosis placement.

**Ultra-mini abstract:** We present the first case series of patient-specific multiscale modeling of the Fontan procedure. Despite noticeable local power loss differences, graft selection does not affect systemic pressure and flow rates or other clinically relevant quantities. Anastomosis placement can affect hepatic distribution.

## Introduction

The staged Fontan procedure remains the common approach to palliate single ventricle circulations. The third stage involves connecting the inferior vena cava and superior vena cava returns to the pulmonary arteries resulting in a pulmonary circulation that is driven by systemic venous pressure only. The most common configuration is the total cavopulmonary connection (TCPC). Although commonly successful in the early to midterm postoperative period, Fontan physiology often worsens as patients mature^1-3^, leading to complications such as arrythmia, protein-losing enteropathy, ventricular disfunction, thromboembolic events, diminished exercise tolerance, fatigue and palpitations^4^. Chronic venous insufficiency, portal hypertension, and retrograde flow in caval veins, which contribute to liver fibrosis or pleural effusions, are also common clinical morbidities for Fontan patients^5^.

In an effort to improve potential flow imbalances to the right and left pulmonary circulations, a Y-graft design was previously developed to replace the extracardiac conduit (ECC) used in the TCPC^6,7^. The earliest Y-graft design was optimized to each patient’s specific anatomy and geometry^6^. The latter Y-graft was chosen using commercially available bifurcating grafts^7^. The promise of the Y-graft was to reduce power loss, thereby reducing ventricular work, and simultaneously to improve distribution of inferior venous flow, which would balance hepatic factors to the right and left lungs^6-8^. Despite the clinical use of the Y-graft in several studies, it remains unclear to what degree these outcomes are achieved and whether power loss of the surgical junction equates to improved performance of the cavopulmonary circulation^9^.

In this study, we constructed patient-specific multiscale models for a cohort of single-ventricle patients undergoing stage 2 to stage 3 surgery. We compared the influence of two Y-graft designs and a traditional extra-cardiac conduit on post-operative outcomes via computational simulation. Patient-specific clinical data was acquired pre-stage 3 and assimilated into physiologic models for each patient in each computational simulation. Multiscale modeling was then used to perform virtual stage 2-3 conversion to compare the performance of graft types. The outcomes of the simulation provide hemodynamic data that is used to assess the physiological consequences of each of the three graft choices.

## Methods

### Patient Selection and Clinical Data

After institutional review board study approval and informed consent for the use of clinical data, six patients were enrolled prior to their pre-operative clinical investigations for planning TCPC. The review entities were Medical University of South Carolina Institutional Review Board and Great Ormond Street Hospital Institutional Review Board. Full approval was given at the start of the project and was renewed yearly until enrollment was completed. Patients were recruited at the Medical University of South Carolina, Charleston, SC, USA, and Great Ormond Street Hospital, London, UK. The pre-operative clinical presentations of the six patients are reported in Table 1. Pre-operative cardiac magnetic resonance imaging (CMR), cardiac catheterization and echocardiography studies were performed prior to surgery in a similar way done for patients studied in Stage 1-2^10^. As in our previous study^10^, pulmonary artery pressure (PAP) was either a direct measurement or an estimate from pulmonary venous wedge pressure. In patients C, E, and F, PAP was acquired on the left side, with no clinical evidence suggestive of a stenosis or cause for discrepancy between the two pulmonary arteries (PAs). In patients A, B, and D, PAPs were acquired on the left and right sides. Only patient D demonstrated anatomical and pressure indications of a left pulmonary artery (LPA) stenosis. Pre-operative echocardiography was performed under GA or sedation. Pulsed wave Doppler traces were acquired in the aorta, SVC, IVC and branch PAs.

**Table 1:**
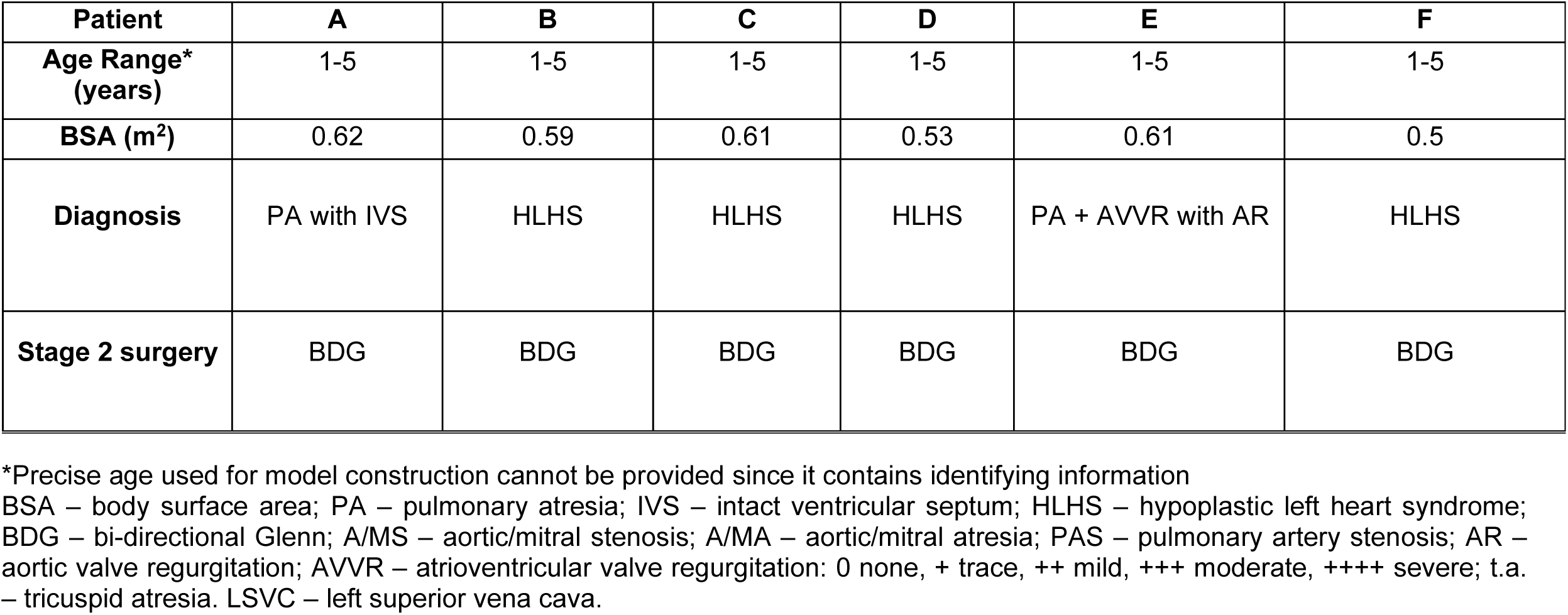
Pre-operative demographics of the six patients used for the study

| **Patient** | **A** | **B** | **C** | **D** | **E** | **F** |
| --- | --- | --- | --- | --- | --- | --- |
| **Age Range\***<br>**(years)** | 1-5 | 1-5 | 1-5 | 1-5 | 1-5 | 1-5 |
| **BSA (m<sup>2</sup>)** | 0.62 | 0.59 | 0.61 | 0.53 | 0.61 | 0.5 |
| **Diagnosis** | PA with IVS | HLHS | HLHS | HLHS | PA + AVVR with AR | HLHS |
| **Stage 2 surgery** | BDG | BDG | BDG | BDG | BDG | BDG |

\*Precise age used for model construction cannot be provided since it contains identifying information
BSA – body surface area; PA – pulmonary atresia; IVS – intact ventricular septum; HLHS – hypoplastic left heart syndrome; BDG – bi-directional Glenn; A/MS – aortic/mitral stenosis; A/MA – aortic/mitral atresia; PAS – pulmonary artery stenosis; AR – aortic valve regurgitation; AVVR – atrioventricular valve regurgitation: 0 none, + trace, ++ mild, +++ moderate, ++++ severe; t.a. – tricuspid atresia. LSVC – left superior vena cava.

### Three-dimensional models and virtual surgery

Three-dimensional models of each patients’ stage 2 anatomy were reconstructed in a similar way as done for Stage 1 patients^10^. Figure 1 depicts the stage 2 to stage 3 reconstructions for the six patients studied ^11,12^. The virtual reconstructed geometry was created following the guidance of a cardiac surgeon, to produce three options of the inferior connections: Y-graft with 12 mm diameter branches (Y 12mm)^6^, Y-graft with 9 mm diameter branches (Y 9mm)^7^, and extracardiac conduit (ECC). The trunk of the graft connecting to the IVC is 18 mm in diameter for all three options. These model designs were based on previously published clinical results from two centers, one of which used a hand-constructed custom design to preserve cross-sectional area from trunk to branches^6^, and the other of which used an “off-the-shelf” readily available design that did not require suture lines at the bifurcation^7^.

**Figure 1:**
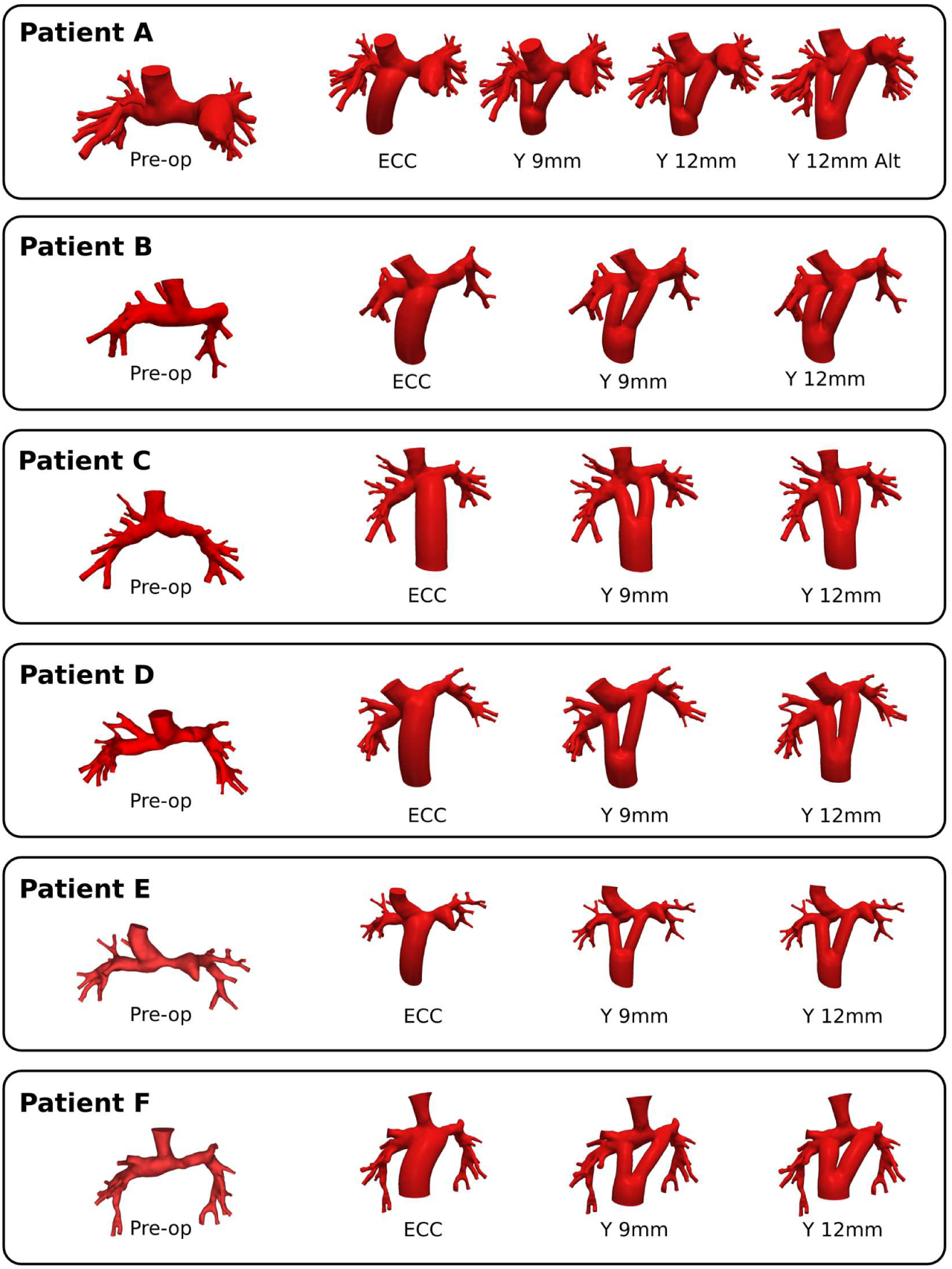
Pre-operative (stage 2) and virtual surgery (stage 3) anatomies for the six patients studied.

### Multiscale Simulation and Analysis

Multi-scale models were developed and tuned for each patient based on the patient-specific anatomical and clinical data (Table 2). Each patient was modeled at the age and body surface area (BSA) at the time of their CMR scan since both 3D and flow information is acquired at this time-point. Following our previous work^11,13^, a 0D lumped-parameter network (LPN) model of the circulatory system outside of the surgical region, was created and coupled directly to the inflow and outflow passages of the 3D model of the surgical site (Figure 2). Previous work ^14^ introduced a method to iteratively tune reduced order (i.e. 3-element Windkessel) representations of the 3D model to match clinical inlet average pressure, inlet average flow and the flow repartition for pulmonary outlets. This was then refined previously^10^ to distinguish between the arterial and venous sides, leading to a 5-element reduced model (Fig S1). At this pre-stage 3 state, flow conservation from clinical data (Table 2) made apparent the presence of collateral vessels that warranted modification of the LPN finally resulting in the model shown in Figure 2. The online supplemental materials detail the inclusion of collaterals in the pulmonary circulation.

**Table 2:**
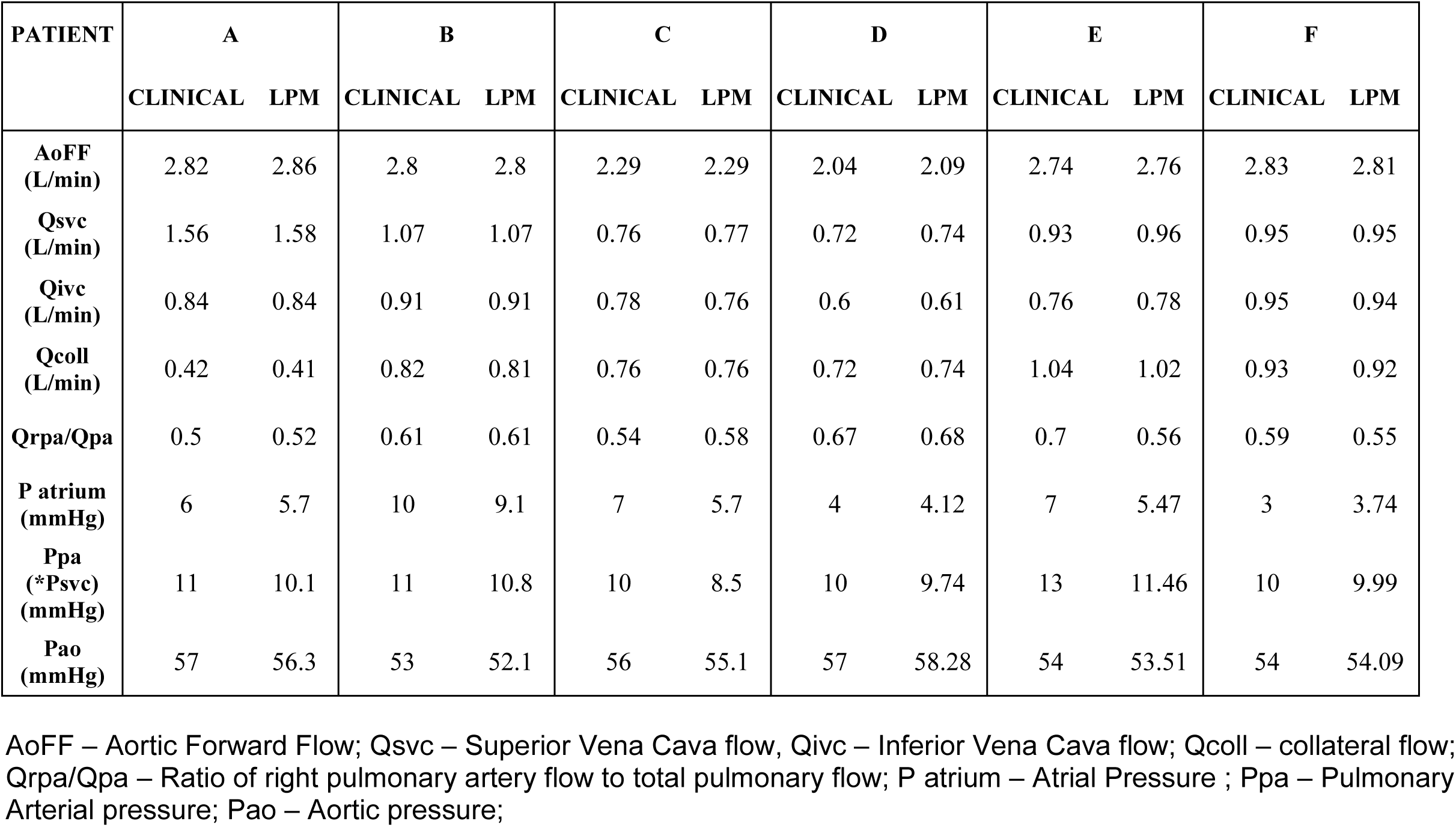
Pre-operative clinical parameters used for patient-specific tuning.

**Figure 2:**
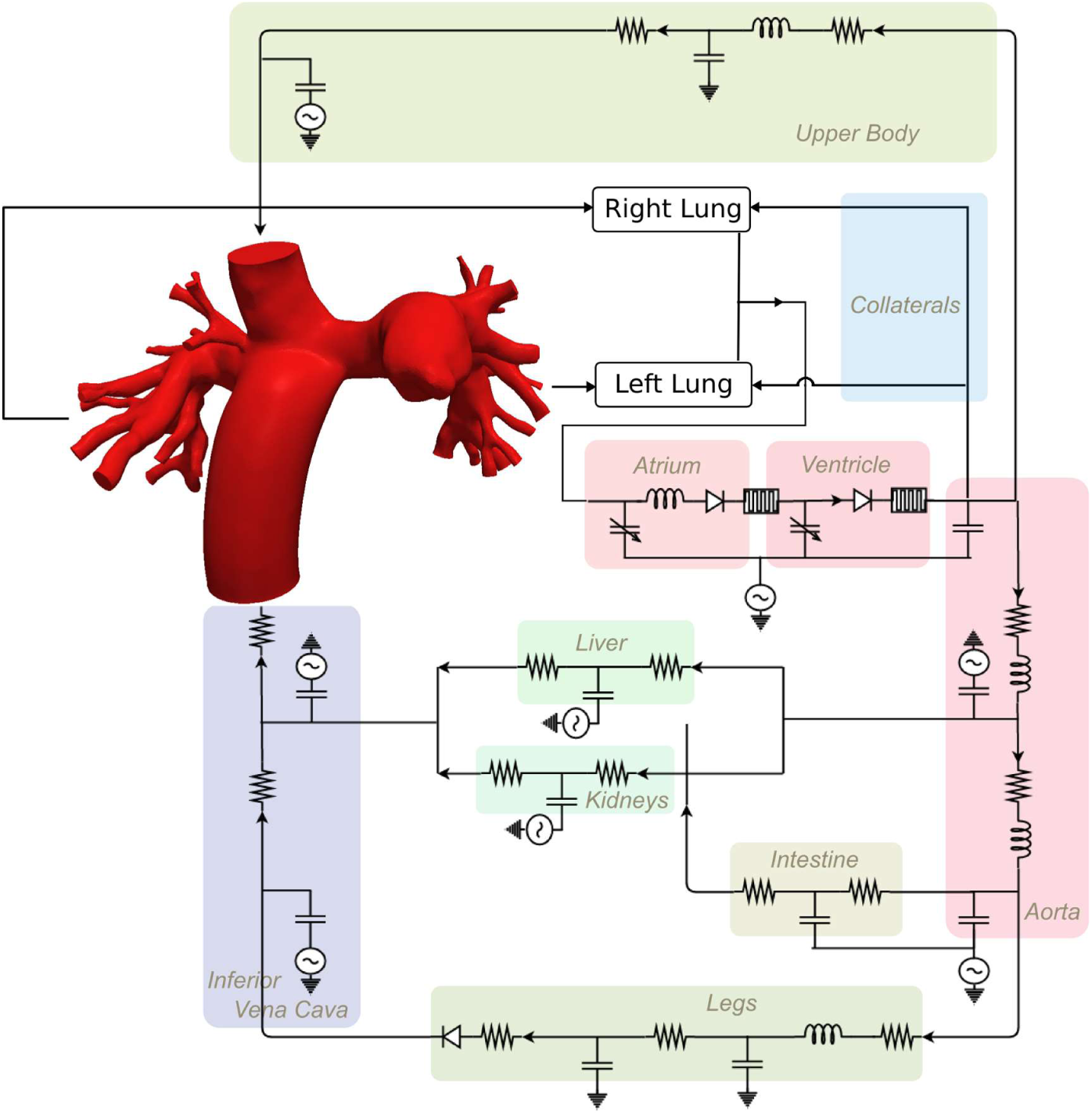
Postoperative multiscale simulation setup.

Multi-scale simulations of the post-operative scenarios were conducted according to previously validated techniques ^11,13,15,16^. Briefly, this involves discretizing the 3D virtual surgery geometries into isotropic finite-element meshes with maximum edge size of 0.03 cm (MESHSIM, Simmetrix Inc., New York) and coupling the 3D Navier-Stokes equations to the 0D LPN using Neumann boundary conditions, implicit coupling, and outflow stabilization ^17^. Flow and pressure in the 3D and LPN domain were solved using a custom incompressible finite element Navier-Stokes solver (Simvascular, www.simtk.org), and a 4^th^ order Runge-Kutta algorithm, respectively. Simulation time step size was 1 ms and 1 μs for the 3D and LPN domain, respectively. Flow and pressure coupling between domains occurs at every 3D-domain time step. Each simulation included 12 cardiac cycles where the last cycle data, by which periodicity had been achieved, was used in the final results analysis. We assume that the walls of the 3D geometrical models are rigid because we do not know the data for the heterogeneous assessment of the wall parameters. Moreover, the anastomosis is stiffened because of sutures.

Power loss was calculated from the simulation results according to our previous publication^13^. Hepatic flow distribution was computed by running an advection solver as a post-processing step. The fluid entering the 3D domain at the IVC outlet face is prescribed to have an “IVC flow concentration” of 1; the advection solver then computes the IVC flow concentration in the entire 3D domain over the cardiac cycle based on flow velocities as the IVC and SVC flows mix. The IVC flow concentration through the left and right PAs is integrated over the cardiac cycle to quantify hepatic flow distribution.

For statistical analyses of the simulation results, we performed one-way ANOVA to quantify the differences between the Fontan graft options. We consider the differences to be statistically insignificant, potentially significant, and significant, when p>0.95, 0.95>p>0.05, and p<0.05, respectively.

## Results

The pre-operative model tuning produced patient-specific results matching clinical data with discrepancies <3.3% for all measured parameters except for the left pulmonary flow ratio and the atrial pressure (where the percentage discrepancies are higher due to the parameters’ values being very small) (Table 2). Notably, The SVC (0.7∼1.6 L/min) and the collateral (0.4∼1 L/min) flow rates are greatly different between the patients, with the standard deviation being approximately 30% of the mean; yet the maximum discrepancy in the tuned model results was only 3.2% and 2.8% for each parameter, respectively (Table 2).

Differences in local hemodynamics between the different graft options are clearly observed in the predicted post-operative flow outcomes. Using results for Patient A as an example (Figure 3), the ECC typically exhibits a direct encroachment and mixing of blood from the IVC and SVC, where the Y-grafts exhibit the SVC flow impacting the inferior PA wall at the anastomosis and the IVC flow merging into the pulmonary arteries. The smaller diameter Y-graft also exhibits higher flow velocities.

**Figure 3:**
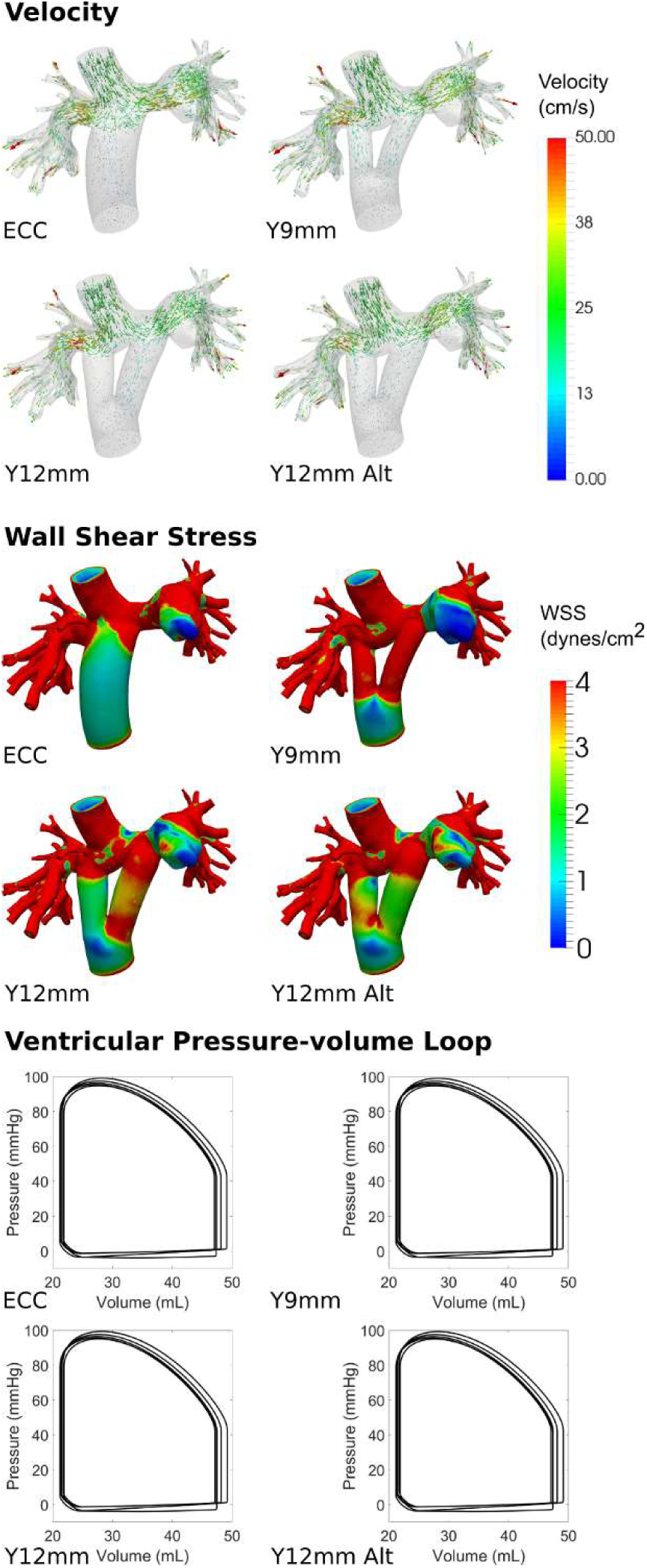
Time-averaged velocity and wall shear stress maps, and ventricular pressure-volume loops for GOSH30.

Between the different graft options, there are noticeable differences (6%∼32%) in the surgical junction power loss for each patient. Due to the direct flow encroachment between the IVC and SVC, the ECC consistently (in 5 out of 6 patients) produced higher power loss compared to the Y-grafts. The power loss in the surgical junction typically contributes 10%∼20% (except in Patient B where it is up to 38%) of the power loss in the total pulmonary circulation (Table 3). The magnitudes of these power losses are compared visually in Figure 4, which reveals that in all patients the maximum power loss difference between graft options are very small relative to the total pulmonary power loss. As a result, in each patient there is negligible difference in the ventricular pressure-volume loop and cardiac work between different graft options (Figure 3). Due to the differences in the total combined cross-sectional area under an unchanging cardiac output, the wall shear stress varies accordingly between different graft options; and depending on the anastomosis positioning, the wall shear stress can be different between the two legs of the Y-graft (Figure 3).

**Table 3:** Post-operative predictions

| PATIENT<br>PROCEDURE | A |  |  |  | B |  |  | C |  |  |
| --- | --- | --- | --- | --- | --- | --- | --- | --- | --- | --- |
|  | Y 9mm | Y 12mm | ECC | Y12 Alt | Y 9mm | Y 12mm | ECC | Y 9mm | Y 12mm | ECC |
| **AoFF**<br>**(L/min)** | 2.77 | 2.77 | 2.76 | 2.77 | 2.84 | 2.84 | 2.84 | 2.29 | 2.29 | 2.29 |
| **Qsvc**<br>**(L/min)** | 1.6 | 1.61 | 1.58 | 1.61 | 1.11 | 1.11 | 1.11 | 0.78 | 0.78 | 0.78 |
| **Qivc (L/min)** | 0.72 | 0.72 | 0.75 | 0.72 | 0.86 | 0.86 | 0.87 | 0.72 | 0.72 | 0.72 |
| **Qcoll (L/min)** | 0.45 | 0.44 | 0.43 | 0.44 | 0.87 | 0.87 | 0.87 | 0.79 | 0.79 | 0.8 |
| **Qrpa/Qpa** | 0.51 | 0.52 | 0.51 | 0.51 | 0.62 | 0.62 | 0.62 | 0.54 | 0.54 | 0.53 |
| **P atrium**<br>**(mmHg)** | 3.02 | 3.04 | 3.01 | 3.03 | 7.1 | 7.11 | 7.12 | 3.18 | 3.18 | 3.17 |
| **Ppa (\*Psvc)**<br>**(mmHg)** | 13.13* | 13.15* | 13.29* | 13.16* | 11.83* | 11.89* | 11.86* | 8.83* | 8.87* | 8.98* |
| **Pao (mmHg)** | 57.42 | 57.48 | 57.47 | 57.45 | 53.13 | 53.15 | 53.14 | 55.24 | 55.26 | 55.26 |
| **Power Loss**<br>**(mW)** | 5.87 | 5.53 | 6.32 | 5.76 | 7.71 | 7.6 | 7.05 | 3.43 | 3.36 | 3.59 |
| **Pa-Sa Loss**<br>**(mW)** | 46.48 | 46.67 | 46.44 | 46.55 | 12.87 | 12.92 | 13.04 | 14.93 | 14.96 | 14.97 |
| **Total Pul Loss** | 52.35 | 52.2 | 52.76 | 52.31 | 20.58 | 20.52 | 20.09 | 18.36 | 18.32 | 18.56 |
| **IVC**<br>**Split Left/All** | 0.49 | 0.66 | 0.33 | 0.49 | 0.39 | 0.36 | 0.34 | 0.42 | 0.36 | 0.61 |

| **PATIENT**<br>**PROCEDURE** | **D** |  |  | **E** |  |  | **F** |  |  |
| --- | --- | --- | --- | --- | --- | --- | --- | --- | --- |
|  | **Y 9mm** | **Y 12mm** | **ECC** | **Y 9mm** | **Y 12mm** | **ECC** | **Y 9mm** | **Y 12mm** | **ECC** |
| **AoFF**<br>**(L/min)** | 2.59 | 2.59 | 2.59 | 3.08 | 3.09 | 3.08 | 3.42 | 3.42 | 3.38 |
| **Qsvc**<br>**(L/min)** | 0.88 | 0.88 | 0.88 | 1.05 | 1.02 | 1.02 | 1.16 | 1.16 | 1.15 |
| **Qivc (L/min)** | 0.63 | 0.63 | 0.63 | 0.82 | 0.86 | 0.85 | 1.04 | 1.05 | 1.02 |
| **Qcoll (L/min)** | 1.08 | 1.08 | 1.08 | 1.21 | 1.21 | 1.22 | 1.21 | 1.21 | 1.21 |
| **Qrpa/Qpa** | 0.61 | 0.61 | 0.6 | 0.66 | 0.66 | 0.66 | 0.57 | 0.56 | 0.59 |
| **P atrium**<br>**(mmHg)** | 5.64 | 5.66 | 5.62 | 6.94 | 6.95 | 6.91 | 3.74 | 3.76 | 3.52 |
| **Ppa (\*Psvc)**<br>**(mmHg)** | 17.83* | 17.82* | 17.92* | 18.7* | 18.74* | 18.86* | 19.02* | 19.06* | 19.31* |
| **Pao (mmHg)** | 73.78 | 73.8 | 73.76 | 62.35 | 62.35 | 62.37 | 72.29 | 72.35 | 71.63 |
| **Power Loss**<br>**(mW)** | 4.4 | 4.17 | 4.83 | 8.8 | 8.8 | 9.46 | 8.02 | 7.85 | 10.62 |
| **Pa-Sa Loss**<br>**(mW)** | 35.03 | 35.11 | 34.9 | 36.34 | 36.33 | 36.19 | 62.54 | 62.66 | 61.28 |
| **Total Pul Loss** | 39.43 | 39.28 | 39.73 | 45.14 | 45.13 | 45.65 | 70.56 | 70.51 | 71.9 |
| **IVC**<br>**Split Left/All** | 0.45 | 0.39 | 0.45 | 0.61 | 0.63 | 0.62 | 0.61 | 0.54 | 0.28 |

AoFF – Aortic Forward Flow; Qsvc – Superior Vena Cava flow Qivc – Inferior Vena Cava flow; Qcoll – collateral flow; Qrpa/Qpa – Ratio of right pulmonary artery flow to total pulmonary flow; P atrium – Atrial Pressure ; Ppa – Pulmonary Arterial pressure; Pao – Aortic pressure

**Figure 4:**
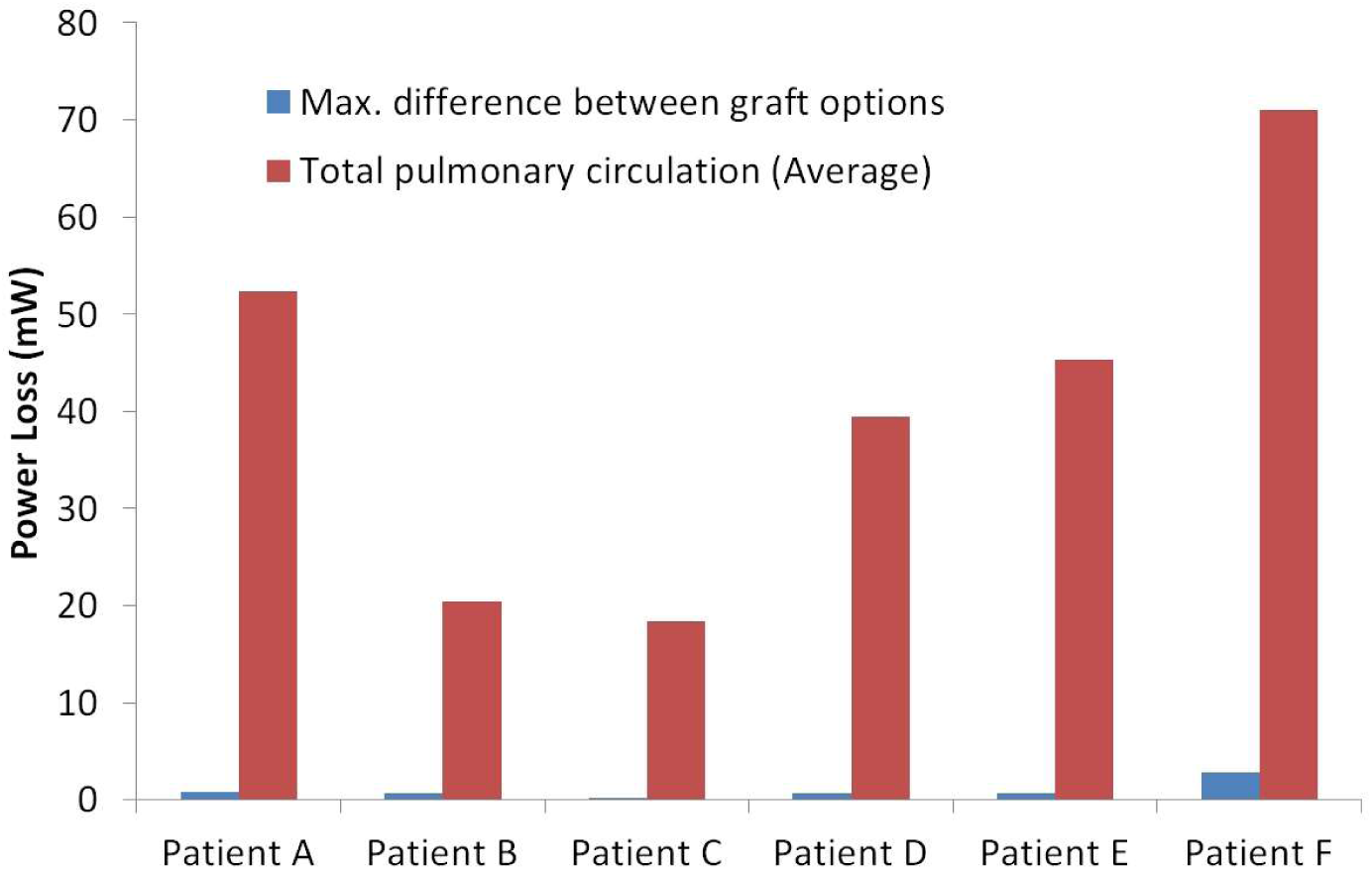
Maximum difference in the local power loss of the Fontan surgical junction between graft options relative to the total power loss in the pulmonary circulation (averaged across all graft option cases).

Statistical analyses revealed insignificant differences between graft options in all physiologic parameters involving pressure and volumetric flow rates (Figure 5). In contrast, there are potentially significant differences in the hepatic flow split between different graft options. This is seen prominently in patient F, where the 9mm Y-graft compared to the ECC produced 61% versus 28% hepatic flow entering the LPA, respectively. Furthermore, it is important to note that changing anastomosis positioning of the same graft option on the same patient can directly affect hepatic distribution; For example, two positionings of the same Y-graft in patient A resulted in 66% versus 49% hepatic flow entering the LPA.

**Figure 5:**
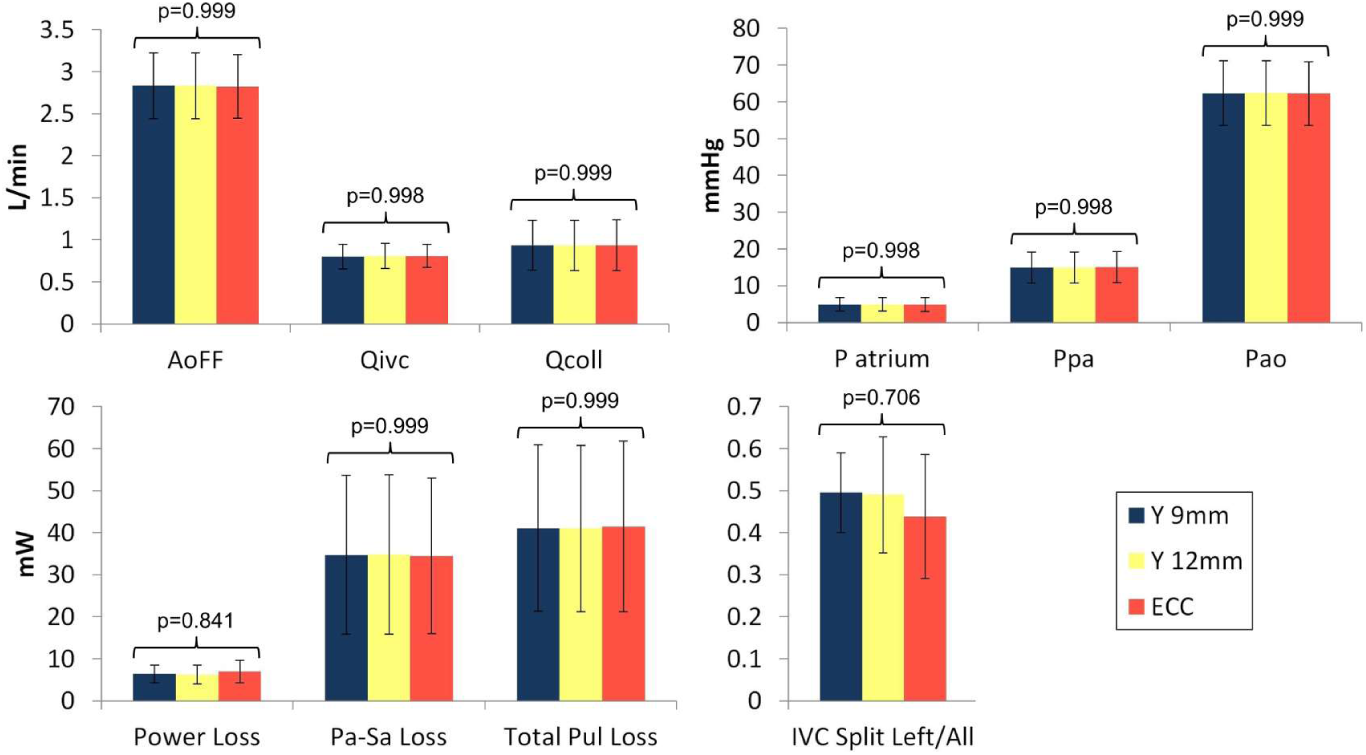
Mean and standard deviation of physiologic parameters compared between graft options, and p-values from one-way ANOVA. AoFF – Aortic Forward Flow; Qivc – Inferior Vena Cava flow; Qcoll – collateral flow; P atrium – Atrial Pressure; Ppa – Pulmonary Arterial pressure; Pao – Aortic pressure

## Discussion

Computational investigations of the Fontan palliations have traditionally focused on the power loss of the isolated surgical junctions. Our multi-scale model results show that differences of greater than 30% in surgical junction power loss still produced negligible effects on most clinically-relevant parameters including cardiac workload and pressure levels in a patient. This finding is consistent with our study on the stage 2 palliation and can be explained by examining the surgical junction power loss in the context of the systemic circuit, an approach possible with our multi-scale computational models. Since much of the power loss of the pulmonary circulation occurs outside of the surgical junction, the patient PVR has a much larger impact on the overall physiology than the hemodynamic differences between different surgical junctions. Figure 4 clearly illustrates that the differences in power loss between different surgical options are very small compared to the total pulmonary power loss in each patient, and thus the impact of graft choice on the overall physiology is minimal. Local flow velocity differences between graft choices, however, may be a consideration factor in term of thrombotic risk. The lower flow velocities in a larger diameter graft may increase thrombotic risk by introducing blood stasis.

Our findings agree with previous single-patient multiscale modeling case studies demonstrating minimal differences in clinically relevant quantifies of interest when changing local geometries in stage 2 and 3 of single ventricle palliation^18^. In particular, similarly small differences in power loss were observed when comparing the hemi-Fontan to bidirectional Glenn surgical choices using multiscale modeling in stage 2 single ventricle patients^19^.

The IVC flow split is an important clinically relevant quantity that is significantly affected by surgical variations. Both graft selection and positioning affect the surgical junction geometry and therefore the resulting IVC flow split. Given the fact that the same graft with different positioning can result in very different IVC flow splits, it is convincing that the actual surgical implementation is the main driving factor in affecting this quantity. Patient-specific multiscale modeling could present considerations for surgical positioning options.

Prior optimization in idealized Y-graft models also showed large differences in hepatic flow distribution when altering anastomosis location and demonstrated that hepatic flow distribution is primarily dominated by IVC / SVC flow ratio and relative LPA and RPA resistances at the model outlets^8,20^. Additionally, this and other studies have demonstrated that hepatic flow distribution is more robust to geometric variations in the Y-graft designs compared to traditional offset designs^21^. Previous studies reporting improvements in hepatic flow distribution with the Y-graft design, compared to the traditional design, are in agreement with the present work^22^.

Despite prior technical success of the Y-graft Fontan in surgical studies, the evidence for implanting the Y-graft solely based on potential to improve energetics of the overall Fontan circulation remains minimal^23,24^. While, improvements to hepatic flow distribution are observed in simulation studies, individual surgical planning should be performed to maximize equal distribution to the pulmonary arteries. It remains unclear whether the increased technical difficulty of the Y-graft procedure is outweighed by the potential benefits of improved hepatic performance, especially in light of the relatively small fraction of Fontan patients who go on to develop arteriovenous malformations (AVMs). However, in certain cases such as patients with interrupted IVC, the Y-graft offers a promising method to reduce the incidence of AVMs, and recent surgical planning studies have demonstrated success in these cases^25,26^.

Modeling simulations provide the opportunity to provide data currently unobtainable by other modalities with essentially no risk to patients. One limitation surrounding modeling is availability of clinical data to ensure validation. The multi-scale models used herein have been validated previously using pre-operative clinical data^10,11,13,14,27^. However, post-surgical data is rarely available and not available for this study. We have previously discussed the type of information required and demonstrate a successful post-surgical validation^28^.

## Conclusion

This paper represents the first case series of patient-specific multiscale modeling of the Fontan procedure. Representations of the patients’ pre-operative physiologies were used to tune the models to match clinical data. Despite noticeable local power loss differences, graft selection does not affect systemic pressure and flow rates or other clinically relevant quantities. However, geometric differences from graft selection or positioning can directly impact IVC flow split, and personalized surgical planning could be used to optimize hepatic flow distribution in these cases.

## Data Availability

NA

## Acknowledgements

This work was supported by the Leducq Foundation through a Transatlantic Network of Excellence Program grant, the National Institute of Health Research Biomedical Research Centre Funding Scheme, a Burroughs Wellcome Fund Career award at the Scientific Interface, an American Heart Association Postdoctoral Fellowship, NSF CAREER OCI-1150184, and a British Heart Foundation Clinical Research Fellowship FS/12/35/29566. We also acknowledge Dr. Charles Taylor, Dr. Nathan Wilson, Mahdi Esmaily, and Weiguang Yang for their contributions to codes used in the simulations and analyses, and the open source Simvascular project at simtk.org.

This report is independent research by the National Institute for Health Research Biomedical Research Centre Funding Scheme. The views expressed in this publication are those of the author(s) and not necessarily those of the NHS, the National Institute for Health Research or the Department of Health.

## Abbreviations

BSA: body surface area
CI: cardiac index
CMR: cardiac magnetic resonance imaging
GA: general anaesthetic
Hb: hemoglobin
HLHS: hypoplastic left heart syndrome
IVC: inferior vena cava
LPA: left pulmonary artery
LPN: lumped parameter network
PAs: pulmonary arteries
PAP: pulmonary artery pressure
PVR: pulmonary vascular resistance
SVC: superior vena cava
TPG: trans-pulmonary gradient

